# Efficacy of federated learning on genomic data: a study on the UK Biobank and the 1000 Genomes Project

**DOI:** 10.1101/2023.01.24.23284898

**Authors:** Dmitry Kolobkov, Satyarth Mishra Sharma, Aleksandr Medvedev, Mikhail Lebedev, Egor Kosaretskiy, Ruslan Vakhitov

## Abstract

Combining training data from multiple sources increases sample size and reduces confounding, leading to more accurate and less biased machine learning models. In healthcare, however, direct pooling of data is often not allowed by data custodians who are accountable for minimizing the exposure of sensitive information. Federated learning offers a promising solution to this problem by training a model in a decentralized manner thus reducing the risks of data leakage. Although there is increasing utilization of federated learning on clinical data, its efficacy on individual-level genomic data has not been studied. This study lays the groundwork for the adoption of federated learning for genomic data by investigating its applicability in two scenarios: phenotype prediction on the UK Biobank data and ancestry prediction on the 1000 Genomes Project data. We show that federated models trained on data split into independent nodes achieve performance close to centralized models, even in the presence of significant inter-node heterogeneity. Additionally, we investigate how federated model accuracy is affected by communication frequency and suggest approaches to reduce computational complexity or communication costs.

## 1 Introduction

### 1.1 Availability of genomic data

The last decade has seen a rapid increase in the amount of genomic data due to the improvement of sequencing technologies and the promise of big data studies in healthcare. With genotyping costs going down, the pool of genomic data also becomes less centralized as more organizations, both commercial, such as genetic testing companies, and non-profit, such as biobanks, accumulate vast collections of genomes. This decentralization, coupled with data-hungry genome-wide machine learning approaches, raises a need for data collaboration. However, access to genomic data is usually restricted due to its sensitive nature and the harmful consequences of possible data leakage, such as deanonymization and genetic discrimination [1] [2] [3].

Data holders may share aggregated data such as summary statistics for genome-wide association studies (GWAS) to allow easy access with a reduced risk of exposing sensitive information. The summary statistics can be analyzed jointly via meta-analysis [4] [5]. However, summarizing involves a loss of information which affects model performance.

Non-profit holders of genomic data, such as biobanks [6] [7], typically allow researchers to access their data upon application approval. While in the case of a successful application, researchers get access to individual-level data, the approval process may take months and joint analysis with other data sources may be prohibited.

### 1.2 Phenotype prediction

Phenotype-from-genotype prediction aims to score an individual’s genetic liability to a certain phenotype, usually, a disease, which can identify risk groups and assist diagnostics [8]. To build a predictive model, one has to obtain either individual-level data where each sample has two alleles for each included genetic variant (SNP) or summary-level data where each SNP has an allele frequency.

Models trained on individual-level data typically yield higher predictive performance as they learn the joint SNP distribution. However, this sensitive data can typically be accessed only with an approved research application.

On the other hand, summary-based models, or polygenic scores, are trained on publicly available GWAS-derived summary statistics and can even incorporate outputs of multiple GWAS using meta-analysis. However, polygenic score models are typically based on assumptions that reduce their applicability to samples with ancestry different from the ancestry of the training set. For instance, multiple studies show poor portability of polygenic scores to other ancestry groups [9] [10] [11]. This is caused by inter-population differences in allele frequency [12] and variant effect size [13], as well as different linkage disequilibrium patterns [14].

In this paper, we consider only individual-level data, since summary-level data is typically publicly available and does not require federated learning to keep it private.

### 1.3 Ancestry prediction

Genetic ancestry prediction from SNPs has two common uses. First, it is a product that genetic testing companies provide to their customers [15]. Despite its not purely scientific purpose, predicted ancestry is an important factor in attracting new customers to provide their DNA samples and, thus, increases the amount of available individual-level genetic data. Second, due to poor polygenic score cross-ancestry portability, per-ancestry summary statistics [16] and polygenic score [17] catalogs have been established. As self-reported ancestry is often noisy, ancestry prediction is a promising tool to be used in phenotype prediction and pharmacogenomics [18].

The task of predicting ancestry is closely related to inferring genetic population structure. As ancestry differences comprise the major part of human genetic variation, population structure is well described by top eigenvectors of the covariance matrix obtained from genetic variants. As a consequence, these principal components are commonly included in phenotype prediction models as covariates to control for population structure [19]. A common approach to estimate the ancestry of unlabeled samples is to project it onto the principal component space of a labeled reference panel [20], such as the 1000 Genomes Project [21].

### 1.4 Federated learning

Federated learning involves training a model locally on clients (data nodes) and sending parameter updates to a server (central hub) where these updates are aggregated into a new set of model parameters which are then sent back to the clients in the next round of training, also called a communication round [22]. Unlike conventional ‘centralized’ machine learning, federated learning does not require assembling data at a single location which saves communication costs and, more importantly, enhances data security since the client’s data is not disclosed to the server nor to other clients.

Federated learning features different strategies, which vary in aggregation methods on a server, in the training process on clients and in the communication frequency between a client and a server. Different strategies may be preferable in different data distribution scenarios: (i) cross-device (many clients with little data) or cross-silo (few clients with a lot of data); (ii) varying degrees of inter-client heterogeneity (dissimilarity); (iii) unavailable or straggling clients. This study considers the cross-silo scenario which is the most typical for genomic data where clients, such as hospitals, biobanks, and genetic testing companies may be dissimilar due to differing genetic populations.

### 1.5 Privacy issues of federated learning

The key privacy-preserving mechanism in federated learning is keeping data in its storage, training a model there and then sending the model to a central server. However, the model may leak information about the data [23, 24] and additional privacy enhancing mechanisms may be required. These include differential privacy [25], secure multiparty computation [26] and trusted execution environments [27], see [1] for a comprehensive survey of privacy-preserving mechanisms.

### 1.6 Federated learning for genomics and healthcare

Researchers have applied federated learning to a variety of healthcare data, including electronic health records [28], medical images [29] and wearables [30]. A number of surveys describe the applications, prospects, challenges and privacy concerns of federated learning in healthcare [31], [32], [33]. A number of privacy-preserving techniques for genomic data have been proposed, such as federated GWAS [34] [35] and federated PCA for GWAS [36]. However, to the best of our knowledge, the efficacy of federated learning for predictions from full-scale genomic data has not been extensively investigated. Training a federated model on genomic arrays poses additional challenges typical to omics data, such as the vast number of non-independent features (genetic variants).

### 1.7 Scope of the paper

In this paper, we analyze the efficacy of federated models trained on individual-level genomic data with the aim to lay the groundwork for the use of federated learning in genomics. First, we show the promise of federated models for phenotype prediction from the large-scale UK Biobank (UKB) genomic data. Next, we analyze federated learning strategies in more detail on the smaller-scale but more heterogeneous 1000 Genomes Project data. We investigate the model behavior on the clients and show the importance of frequent communication to achieve faster convergence in the presence of significant heterogeneity (client dissimilarity). In both experiments, we use well-established and accurate models and train them using the standard FedAvg strategy on artificially-separated datasets to provide a baseline and focus on the analysis of federated model behavior. We also do not consider additional privacy-enhancing mechanisms. Future studies may focus on using FL with more advanced models, designing novel FL strategies, protecting federated models against the attacks, as well as train federated models on multiple real datasets, such as biobanks.

## 2 Results

### 2.1 Phenotype prediction from UK Biobank data

In this experiment, we mimic the situation where genomic data is stored in multiple large silos within the same country. We split the UK Biobank data into 19 datasets according to sample collection centers in different parts of the UK. Some inter-node heterogeneity is present due to the correlation between the UK’s genetic population structure and geography [37]. After a standard quality control (QC), we reduced dimensionality by conducting GWAS on each node and selecting top SNPs. Then, we trained local, federated and centralized Lassonet neural networks (see Methods) of identical architecture with selected SNPs, sex and age as features. Advanced lasso-based models are considered to be state-of-the-art for phenotype prediction from individual-level genomic data [38, 39]. Here, we chose a basic Lasso-based model to focus on the relative performance of federated, centralized and local models. The experiment setup is visualized in Figure 1 and described in more detail in the Methods section.

**Figure 1:**
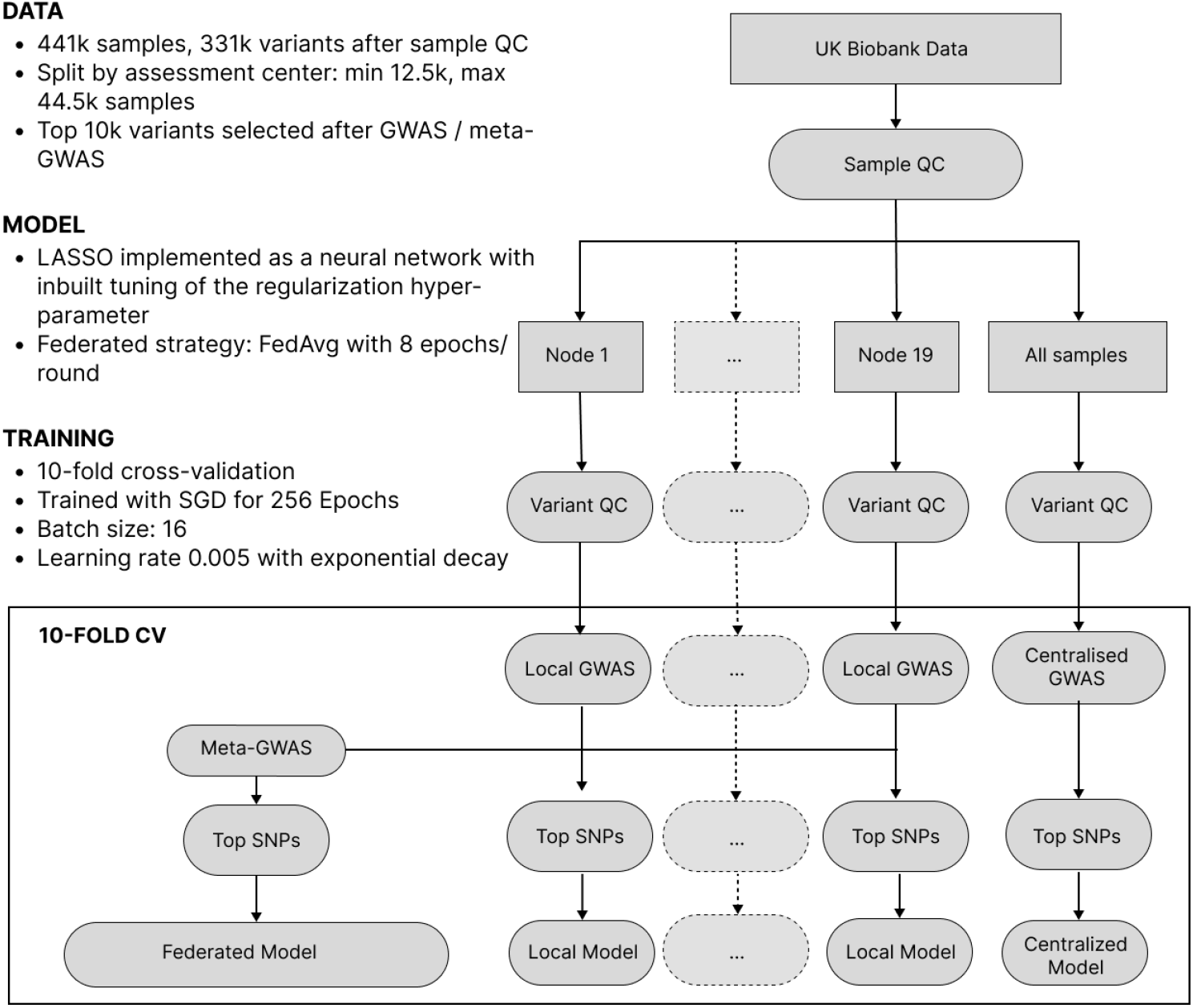
The workflow of phenotype prediction from UKB data. Data is split into nodes and undergoes per-node variant QC. Then, within a cross-validation loop dimensionality is reduced by selecting the most significant SNPs via a local or meta-GWAS. Finally, models are trained.

Figure 2 displays test *R*^2^ performance of six out of 19 local models, federated (FedAvg, 8 epochs in a communication round, see Methods) and centralized models for nine continuous phenotypes. The six displayed nodes were selected, prior to model training, to demonstrate the whole range of sample sizes. The considered phenotypes were selected based on previous heritability estimates [40]. On each node two local models were trained: one trained on top SNPs from local GWAS and one trained on top SNPs from meta-GWAS (see Methods). The federated models used SNPs from meta-GWAS and the centralized models used SNPs from centralized GWAS. Local and centralized covariates-only (sex and age) models were also trained as a baseline.

**Figure 2:**
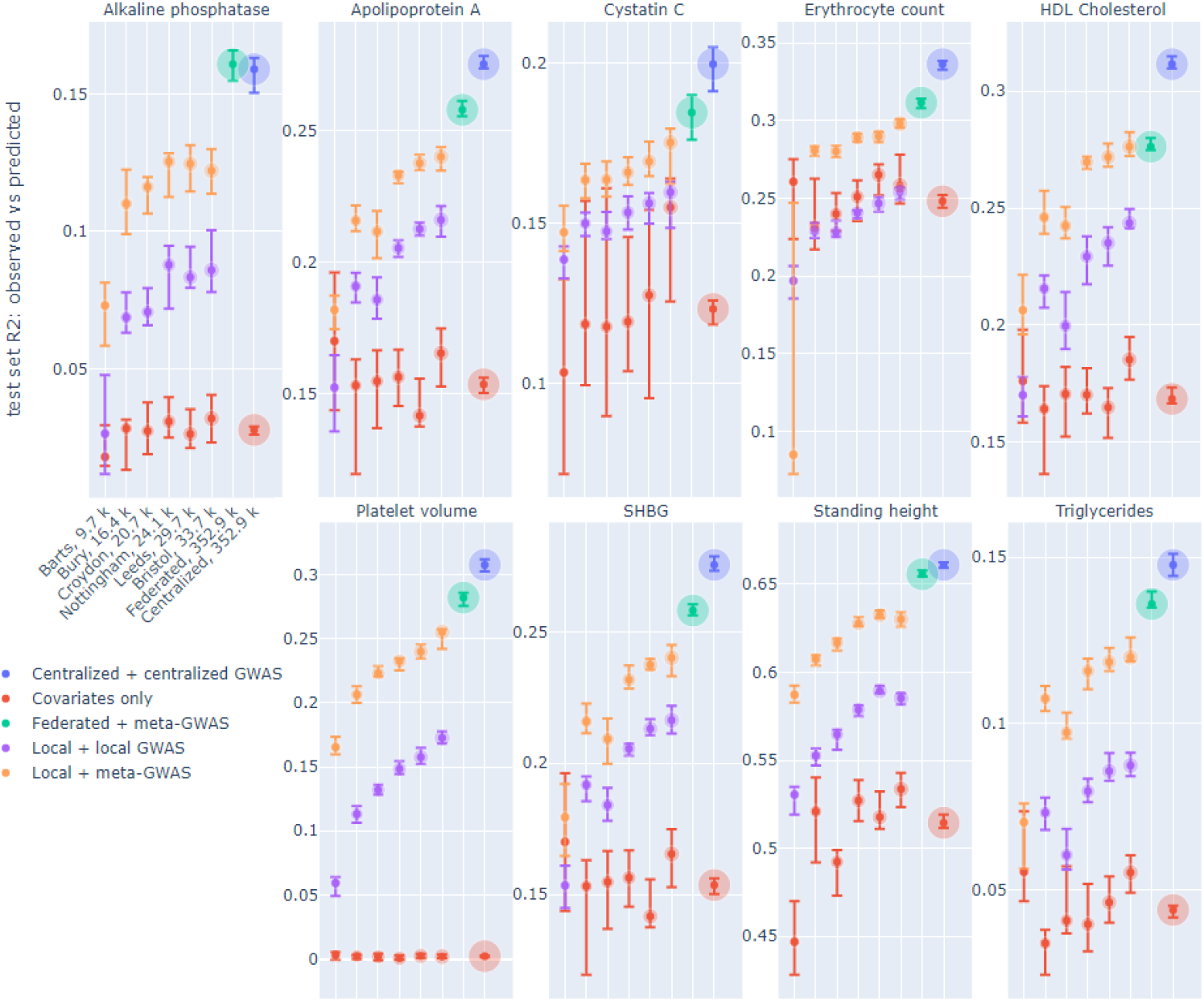
*R*^2^ of observed vs predicted phenotype on a test set of local, federated and centralized models for phenotype prediction on UK Biobank data split by assessment center. Facets correspond to the phenotypes. The x-axis shows six of 19 local nodes, selected to represent a range of node sizes, in increasing size order, which is the same for each phenotype. The number of training samples in each node is indicated in the x-axis labels, which bubble size also corresponds to. Color corresponds to the model type. For each dot, the median and 80% confidence interval for 10-fold cross-validation is displayed.

We trained local and centralized models with (i) ‘native’ features, i.e. SNPs derived from local and centralized GWAS, to represent end-to-end solutions, and (ii) SNPs yielded by meta-GWAS so that local, centralized and federated models can be compared on exactly the same set of features. For centralized models, performance on centralized GWAS SNPs and meta-GWAS SNPs was very similar for all phenotypes, thus, only the former was included in the Figure 2. Local models tend to perform better on meta-GWAS SNPs compared to local GWAS SNPs, most likely because SNP selection via a local GWAS tends to yield more false positives due to an insufficient number of samples. For local models, we see a natural trend that performance improves as the node size grows. Federated models outperform all local models and get close to centralized models.

### 2.2 Ancestry prediction from 1000 Genomes data

Despite its lower clinical significance than predicting complex traits from genotype, we now consider ancestry-from-genotype prediction models. They can be trained using a smaller number of samples and are lighter and less computationally expensive, which allows us to conduct extensive simulations to investigate federated learning in more detail. Here, we mimic the situation where genetic testing companies from different parts of the world collaboratively predict ancestry and split the 1000 Genomes Project data into 5 isolated nodes based on sample superpopulation (African, Native American, East Asian, European, Southern Asian), thus getting high inter-node heterogeneity. After a standard QC, we reduced dimensionality by applying federated PCA [41] to pruned SNPs and then trained local, federated and centralized multilayer perceptrons (MLPs) of identical architecture. The experiment setup is visualized in Figure 3 and described in more detail in the Methods section.

**Figure 3:**
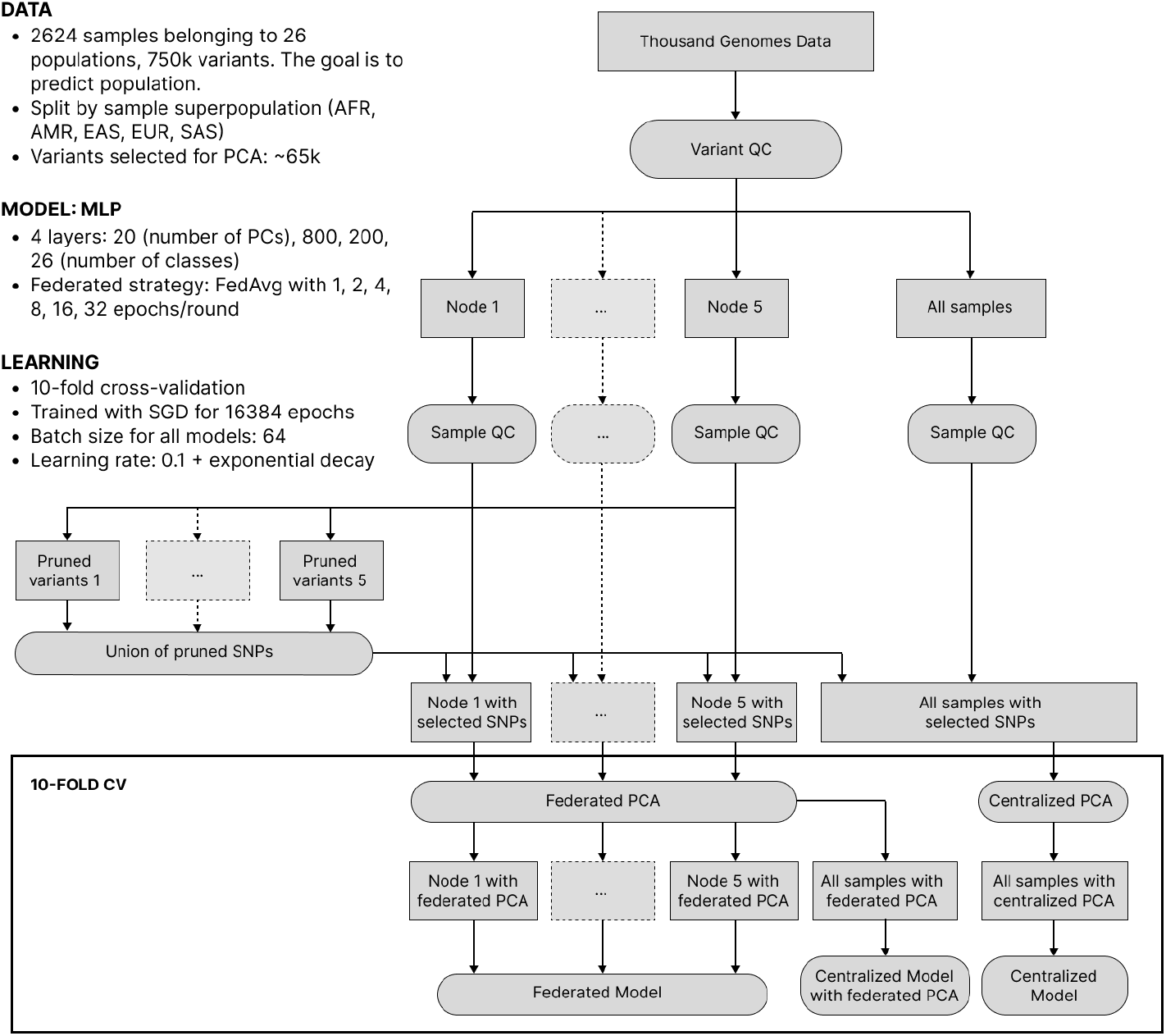
The workflow of ancestry prediction from 1000 Genomes data. Data was split into nodes according to sample superpopulation. The union of variants pruned on each node was taken for all nodes to have the same features. Federated / centralized PCA was used to further decrease dimensionality. Finally, federated and centralized models were trained.

Our goal here is to investigate the performance of federated models as a function of communication between the clients (nodes) and the server in the presence of significant cross-client heterogeneity. For this, we compare FedAvg strategies with a different number of epochs in a round of communication. The training process of federated models is displayed in Figure 4.

**Figure 4:**
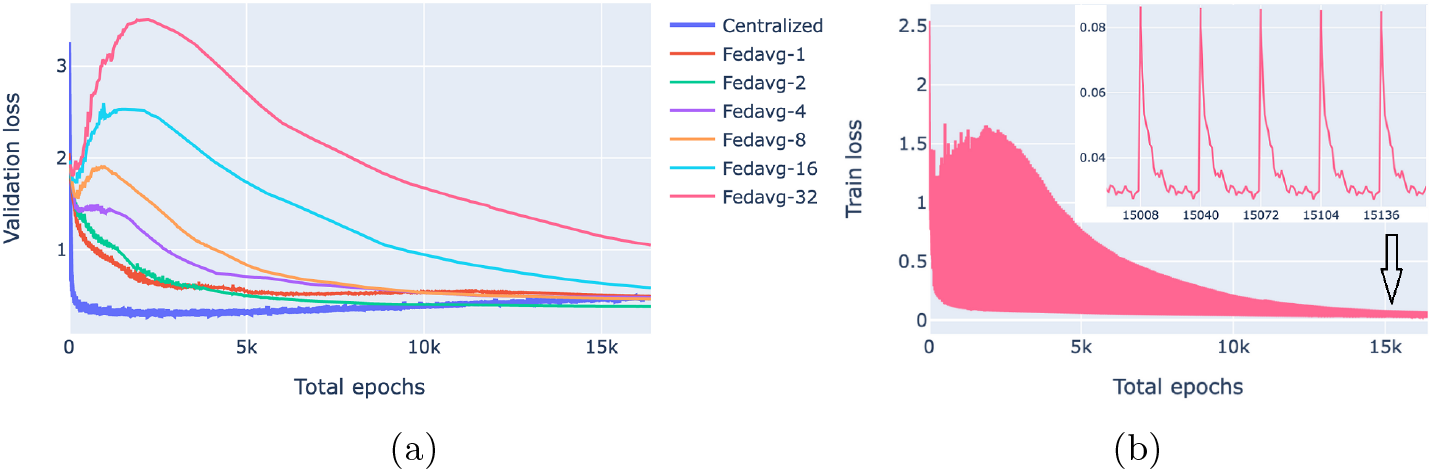
Loss behavior of federated and centralized models. FedAvg strategy with 1, 2, 4, 8, 16, 32 epochs/round was used. (a) Validation loss of federated models with a different number of communication rounds. For each model and epoch, a median loss over 10-fold cross-validation is shown. (b) The training process of a 32-epochs-in-round model on a client. Every 32nd epoch contains two points: one before the start of the epoch when the client receives parameters from the server and one after the first epoch of a round.

Figure 4a compares the validation loss of the centralized and federated models. Federated models show a clear trend that the more communication between the server and the clients, i.e. the more rounds and the fewer local epochs in each round, the faster the convergence is. For the centralized model, convergence was fast, however, it started from a higher loss value because of the random class assignment in a multiclass classification initialization: a centralized model solves a problem for 26 classes whereas each of the five superpopulation nodes has fewer classes (26 in total).

Figure 4b shows the evolution of the client training loss of the FedAvg model with 32 local epochs in a round of communication. Here, the peaks correspond to the initial evaluation of the aggregated parameters sent from the server to the client. Parameter aggregation on the server results in an increase of the local loss as the aggregated parameters are a weighted average of parameters optimized on different data distributions (due to inter-node heterogeneity), then the loss starts decreasing as the model starts fitting to the local data.

### 2.3 Practical considerations of server-client communication

Figure 5 shows the accuracy of federated models as functions of the number of total epochs (computational complexity) and rounds (communication). In compliance with Figure 4a, Figure 5a shows that for heterogeneous data, increasing communication between the clients and the server leads to higher accuracy. On the other hand, Figure 5b shows that for a limited number of rounds, it is beneficial to train locally for a larger number of epochs. Thus, depending on what is the bottleneck of the system, communication or computational complexity, different federated learning strategies may be preferable.

**Figure 5:**
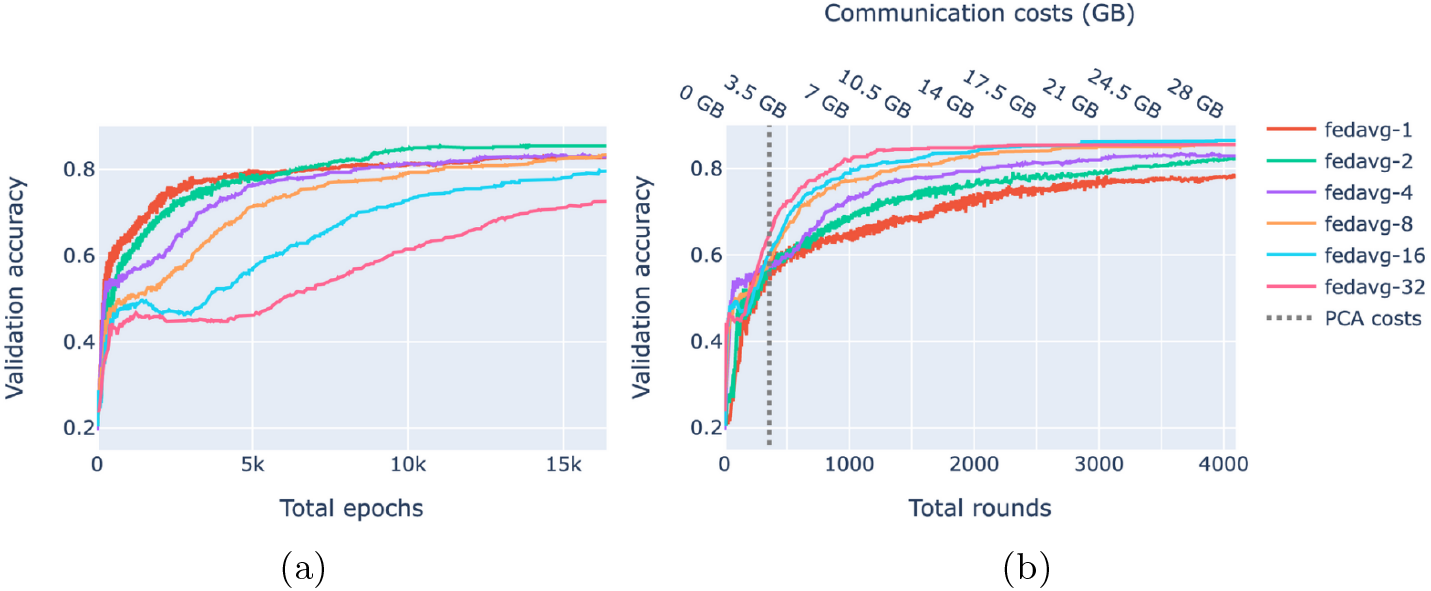
Validation accuracy as a function of complexity and communication. FedAvg strategy with 1, 2, 4, 8, 16, 32 epochs/round was used. Each shown value is a median over 10-fold cross-validation. (a) Accuracy of federated models as a function of the total number of epochs. (b) Accuracy of federated models as a function of the amount of communication between the server and the clients. The dashed line corresponds to the amount of communication used by federated PCA.

A fully federated solution requires all data to be prepared in a federated manner as well. In the case of ancestry prediction, dimensionality reduction is usually conducted via PCA, thus, we first pruned SNPs as displayed in Figure 3 to decrease computational load and then utilized federated PCA using the P-stack algorithm as described in [41]. The amount of communication used in federated PCA linearly depends on the number of input SNPs which affects the model accuracy. Hence, if communication is limited, one can spend more on the PCA step by including more SNPs or use more communication rounds for model training. Figure 6 shows our rationale behind choosing the pruning parameters that determine the number of input SNPs for federated PCA. We also validate the federated PCA approach and show that the centralized classifier performs similarly on federated and centralized PCs.

**Figure 6:**
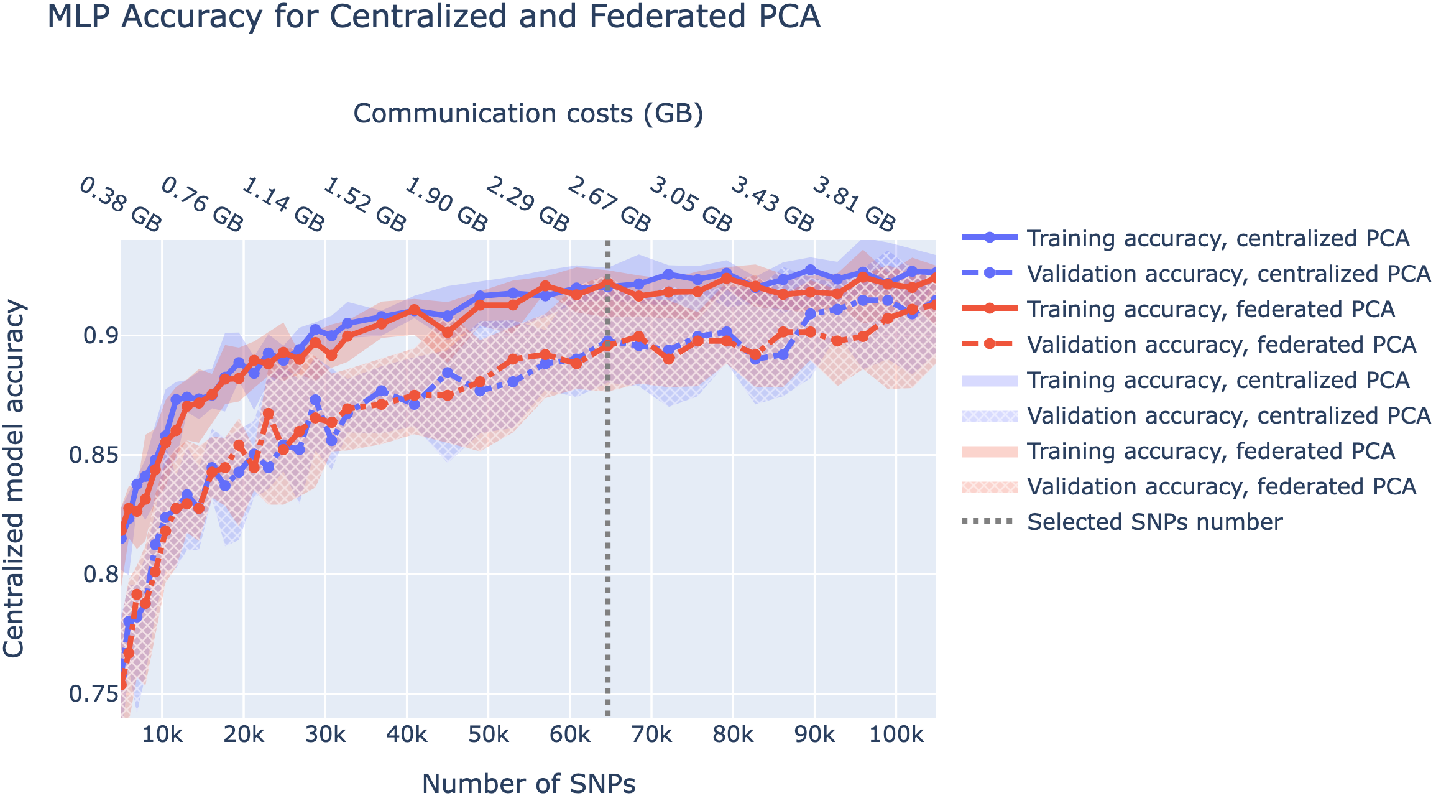
Centralized model accuracy as a function of the number of SNPs used for dimensionality reduction via centralized and federated PCA. Solid/dashed line corresponds to train/validation model accuracy, and blue/red corresponds to centralized/federated PCA. Shaded areas correspond to 80% confidence intervals based on 10-fold cross-validation. Vertical dashed lines correspond to the number of SNPs we chose to be used in downstream analysis.

## 3 Methods

This study aims to assess the applicability of federated learning to genomic data. In this paper, we consider only ‘global’ models that aim to perform well on all varieties of data split between isolated datasets. A survey of personalization methods for federated models is provided in [42].

Intuitively, we expect a good federated model to perform considerably better than the local models, trained on data in a single node, and slightly worse than a centralized model that trains on all of the data together. In this case, a federated model delivers all benefits of federated learning at the cost of a small reduction in performance compared to a centralized model.

### 3.1 Federated learning strategies

Here, we consider the FedAvg strategy [43] with a different number of epochs in a communication round. In our case, where the number of clients is low, the pseudocode is displayed in Algorithm 1:

#### Algorithm 1

FedAvg for a small number of clients. *K* is number of clients, *n*_*k*_ – number of samples on *k*th client, *R* – number of communication rounds, *E* – number of local epochs in a round, *η* – local learning rate

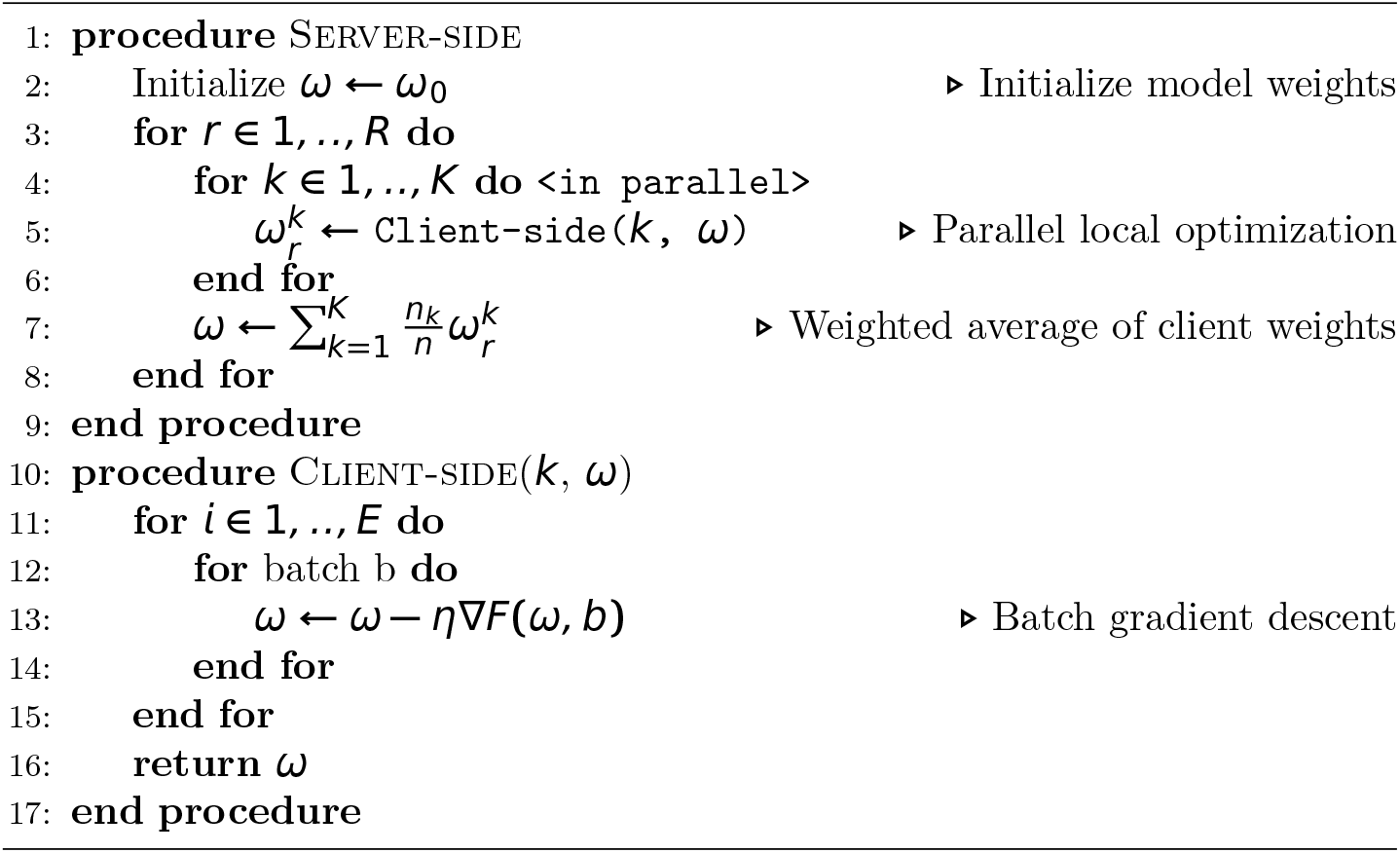

In the presence of significant inter-node heterogeneity, i.e. when the local data distribution on the clients is different from the global distribution, the global model tends to overfit to local data causing ‘client drift’ that slows or prevents convergence [44]. Client drift can be decreased by limiting the amount of training on a client in a single round, i.e. increasing communication between a client and a server. In this paper, we utilize FedAvg with different amounts of server-client communication by varying the number of communication rounds *K* but keeping the total number of local epochs *KE* constant.

### 3.2 The UK Biobank dataset and data processing

An overview of our data processing pipeline for experiments on the UK Biobank dataset can be seen in Figure 1. All samples underwent quality control using PLINK [45] to remove individuals with an insufficient number of genotyped variants (6% missingness cutoff) and related samples with a KING [46] cutoff 0.0884 corresponding to second-degree relatives. For local and federated but not centralized models, the data is split into 19 datasets according to the data collection center (UKB data-field 54). Data collection centers with less than 10k samples were excluded. The size of the datasets after QC ranges from 12.1k (Barts) to 42.1k (Bristol), for a total of 441k samples. For each dataset separately, variant QC is conducted by filtering out variants by rarity (5% minor allele fraction cutoff), missingness (2% cutoff), and a Hardy-Weinberg equilibrium p-value threshold of 10e-6. Each dataset is split into 10 folds for cross-validation where at each time eight folds are used as training data, one as validation data (to be used for early stopping during model training and regularization parameter selection), and one as test data. Next, we reduce dimensionality by selecting the 10,000 most significant SNPs with a GWAS conducted on the training data using age, sex, and 20 principal components from SNPs as covariates. These 10,000 SNPs are combined with age and sex as the input features for our predictive model. For experiments with federated models, feature selection is done by performing a random-effects meta-analysis using PLINK to aggregate information from the GWAS reports of the individual datasets and then selecting the top 10,000 SNPs.

### 3.3 Phenotype prediction model

The best-performing phenotype prediction models on large datasets like UK Biobank are typically LASSO-based and are solved iteratively to save memory consumption [47, 38, 39]. LASSO is a linear model with L1 regularization, which performs feature selection by setting the weights of non-influential parameters to zero. A LASSO problem can be solved using coordinate descent [47] or gradient descent. We chose to solve LASSO using gradient descent because it can be easily implemented on top of existing deep learning and federated learning frameworks, such as PyTorch [48] and Flower [49].

However, with gradient descent, the LASSO problem can only be solved for a single value of the regularization parameter *λ*. Since *λ* determines model performance and generalization ability, one typically trains multiple models with different values of *λ* and selects the one with the best validation metric. We implemented this procedure as a linear neural network which efficiently trains LASSO models with a range of *λ* values in parallel and offers built-in model selection. We call this implementation Lassonet.

Centralized and local Lassonet models are trained using PyTorch and PyTorch Lightning [50] on the Zhores cluster node with Nvidia V100 GPU with 16GB VRAM and up to 160GB of RAM [51]. We used the SGD optimizer with learning rate 5e-3, learning rate decay 0.99, batch size 16 and trained Lassonet for 256 local epochs for each run on the UK Biobank data.

We implemented federated models using the Flower framework. Here, we aggregated validation loss from models with the same *λ* across clients each round, and then chose the model with the best validation loss to be evaluated on the test set.

### 3.4 The 1000 Genomes Project dataset and data processing

The 1000 Genomes array contains about 750 thousand genetic variants (SNPs) of 2624 samples of 26 genetic populations belonging to five superpopulations of East Asians (EAS), Southern Asians (SAS), Europeans (EUR), Africans (AFR) and Native Americans (AMR).

Our data processing workflow sketched in Figure 3 was performed in the following order. First, we conduct variant QC in PLINK keeping genetic variants with minor allele frequency *>*5% and missing call rates *<*2%. Next, we split the samples into five isolated nodes according to sample superpopulations. Then, we conduct sample QC on each node separately in PLINK keeping non-related (KING relatedness cutoff 0.0884 that corresponds to second-degree relatives) samples with missing call rates *<*6%.

Next, we reduced dimensionality by first pruning variants on each node separately in PLINK, then taking a union of the remaining variants across the nodes to get a single variant set (only variant IDs are communicated between nodes) yielding about 65 thousand SNPs. Then, data on each node was split into 10 folds for cross-validation where at each time eight folds are used as training data, one as validation data (to be used for early stopping during model training) and one as test data. Finally, we conducted federated PCA and, alternatively, centralized PCA on the training set and extracted the top 20 principal components. To reduce the dimensionality of validation and test sets, we projected them onto the training PC space. The influence of the pruning strictness and the federated vs centralized PCA is displayed in Figure 6.

### 3.5 Federated PCA for dimensionality reduction

The standard way to reduce dimensionality for ancestry-from-genotype prediction is by using the principal component transformation as it is well known that PCs of genetic variants retain genetic population structure in the dataset [52]. The federated models we use in this study require client datasets to have the same feature space, therefore the PCs have to be obtained collaboratively. Since computing PCs centrally discloses the data and therefore compromizes the purpose of downstream federated learning, we employ the federated PCA approach.

We utilized the P-STACK method as described in [41], which involves sending a local eigenvalues vector and an eigenvector matrix from each client to the server. Further, the server stacks local PCA components and then performs a singular value decomposition (SVD) of the obtained joint matrix. To perform an exact PCA, we used the maximum available number of eigenvectors on each client, which equals the number of client samples minus one. When the number of PCs is fixed, the size of the eigenvector matrix depends only on the number of genetic variants. We used PLINK to prune genetic variants, i.e. removed variants in close linkage disequilibrium. Pruning was conducted on each individual node and then a union of remaining on each node variants was taken.

This allowed us to reduce communication costs for federated PCA and significantly shrink RAM consumption while running SVD on the server. When SVD is completed, the resulting eigenvector matrix is sent back to the clients after which each client is able to perform the PC transformation into the joint feature space.

### 3.6 Ancestry prediction model

After performing the Federated PCA on the 1000 Genomes dataset we use 20 PCs as features for a multi-layer perceptron neural network. It has two hidden layers and outputs raw scores of a sample belonging to the each of 26 populations. It has approximately 182K parameters.

We use a fully-connected neural network with two hidden layers of size 800 and 200, respectively, 20 input and 26 output neurons. The activation function is *selu* and the loss function is cross entropy. We trained it for 16384 local epochs with batch size 64, learning rate 0.1 and exponential learning rate decay with *γ* 0.9999. We trained it on CPU-only machines with 4 CPUs and 8-16 GB of RAM.

## 4 Discussion

The promise of federated learning for healthcare, and genomics in particular, is a result of two powerful trends. First, machine learning models require a lot of data to train and their applicability depends on the diversity of the training dataset. Training the model on diverse data obtained from multiple sources reduces confounding by population genetics, experimental design, etc. and generally improves performance on external data. Second, the growing awareness of the sensitivity of healthcare data and the harmful consequences of its leakage encourages data custodians to restrict data access, e.g. by requiring an application approval and then granting access only within a trusted research environment [53] [54]. This makes merging multiple datasets at a single location challenging, thus discouraging training conventional centralized models.

Nevertheless, the applicability of federated learning to individual-level genomic data has not been studied extensively. In this paper, we analyzed the behavior of federated models in two scenarios: phenotype-from-genotype prediction on the UK Biobank data and ancestry-from-genotype prediction on the 1000 Genomes Project data. We first showed that federated models are almost as accurate as centralized models and considerably more accurate than local models for predicting multiple phenotypes from genomic data.

It would have been interesting to split UKB to nodes by ethnic background and see how FL performs in the presence of higher node heterogeneity. However, non-European nodes in UKB would have less than 10000 samples which may not be enough to make robust predictions of complex phenotypes. On the other hand, even hundreds of samples are enough to make accurate ancestry predictions, due to the fact that ancestry has much more genetic variation than complex phenotypes. Therefore, we moved to ancestry prediction using the 1000 Genomes dataset which features fewer samples but higher population diversity. By splitting the data by sample superpopulations we achieved high inter-node heterogeneity. We showed that in this setting, frequent communication between the server and the clients plays a crucial role in achieving fast convergence and showing performance similar to that of the centralized model. We also demonstrated that depending on whether computational time or communication is a bottleneck of the system, FedAvg with different numbers of epochs in a round should be preferred.

In both of our experiments, the main reason for federated models not reaching the performance level of centralized models is data heterogeneity across the nodes or client dissimilarity. When a federated model trains on a client, it over-fits to local data; then as fitted parameters from different clients get aggregated, the result may differ from the update of the corresponding centralized model, a phenomenon called client drift. Client drift can be decreased by increasing communication between the client and the server by decreasing the number of epochs of local training in a communication round (between parameter updates), as shown in Figures 4a and 5a.

Federated learning is a quickly developing field of research and new strategies continue to emerge, including those aiming to tackle client drift and improve convergence in case of high inter-node heterogeneity, such as SCAFFOLD [55] and FedDyn [56]. These novel strategies may be preferable if communication between the clients and the server is limited. However, they require additional testing, as FedAvg with frequent communication, one or two epochs in a round, is a difficult baseline to beat.

In both experiments, we used a single dataset artificially split into several independent nodes. On the one hand, this is an advantage as the uniform data collection process allows us to limit the influence of environmental and experimental confounders and focus on the relative performance of the models. On the other hand, in a real scenario, using multiple independently collected datasets may require additional work to unify features and outputs across datasets. For example, predictions from genomic data may require SNP imputation or another solution if different datasets have different sets of genetic variants; similarly, ancestry and phenotypes may be defined or collected differently in different experiments.

Federated learning enables data collaboration in genomics which may help solve several important problems. First, combining multiple datasets increases sample size which improves overall model accuracy and enables the prediction of rare diseases and inclusion of rare variants, which typically have larger effect sizes [57]. Second, the vast majority of healthcare data currently comes from people of European descent, which makes models trained on this data biased towards Europeans, adding to the healthcare inequality of people around the world [58] [59]. Federated learning allows to include smaller datasets of different ancestries in the analysis and, thus, reduce the bias.

Being one of the first papers to explore federated learning on genomic data, this study has a limited scope. First, here we assume the trustworthiness of the parties. This is a reasonable assumption if data custodians, such as biobanks, give access to data upon application approval. However, data collaboration between different entities may require implementing additional privacy-enhancing mechanisms, as federated learning does not fully protect against privacy leakage. Second, we used basic machine learning models and the standard FedAvg strategy to keep things simple and focus on the relative performance of federated vs centralized vs local models. Depending on a specific problem, fine-tuned models may yield higher absolute performance; other FL strategies may achieve faster convergence with less communication. Third, as mentioned previously, establishing a real data collaboration may involve additional work to harmonize features and outputs between parties. Fourth, here we focused only on building a single ‘global’ model, whereas some parties in data collaboration may require ‘personalized’ models that prioritize their data [42]. We hope that these issues will be addressed in future studies.

## 5 Conclusions

Despite its promise, the applicability of federated learning to individual-level genomic data has not been sufficiently investigated. We filled this gap by training and analyzing federated models in two important scenarios: phenotype-from-genotype prediction and ancestry-from-genotype prediction. We showed that federated models consistently achieve high performance close to that of centralized models for the prediction of multiple phenotypes and ancestry, even in the presence of significant inter-node heterogeneity. For heterogeneous nodes, we investigated the dependency of federated models convergence on the amount of communication between the server and the nodes and provided recommendations on which schedule to choose if communication or computational time is a bottleneck. We also showed how federated prediction models can be integrated with federated data processing steps such as dimensionality reduction by federated PCA. This study encourages the adoption of federated models in healthcare, which has the potential to enable global data collaboration and train less biased models that represent diverse genetic ancestries.

## Data Availability

All data produced in the present study are available upon reasonable request to the authors.
UK Biobank data are available upon request through the UK Biobank website.

## 6 Acknowledgements and data availability

The computations performed in this study on the UK Biobank dataset were done on the Zhores cluster [51], and we thank the CDISE HPC team for their assistance. This research has been conducted using the UK Biobank Resource under Application Number ‘43661’. We also thank Dmitry Yarotsky, Pavel Nikonorov and Bert Mouler for the valuable discussions. All code written in support of this paper is publicly available at https://github.com/genxnetwork/fl-genomics. The study was funded by GENXT LTD.

## Notes

### Competing Interest Statement

All authors had financial support from GENXT LTD for the submitted work.

### Funding Statement

The study was funded by GENXT LTD, no external funding was received.

### Author Declarations

Access to UK Biobank data was granted upon application 43661. UK Biobank has approval from the North West Multi-centre Research Ethics Committee (MREC) to obtain and disseminate data and samples from the participants (http://www.ukbiobank.ac.uk/ethics/), and these ethical regulations cover the work in this study. Written informed consent was obtained from all participants.

### Summary of Updates

Updated some claims and wording.

